# Social vs individual training: a cross-sectional observational study of perceived motivation, safety, effort, mood, and satisfaction in recreational exercisers

**DOI:** 10.64898/2026.07.23.26358772

**Authors:** Sorrentino Marco, Prof.ssa Castenetto Michelle, Cantù Paolo, ssa Landenna Alessia, ssa Zakar Alice, Inglese Federico, Botturi Filippo, ssa Cozzi Francesca, Profera Giacomo, ssa Combi Lucia, ssa Angeletti Valeria, Vellutata Vito

## Abstract

**Background:** Training context may shape motivation, perceived effort, confidence, mood, and satisfaction during exercise, but evidence from real-world self-reported samples remains limited. This dataset includes 155 respondents and compares individuals who reported training mainly alone with those who reported training mainly in groups.

**Objective:** To describe the sample, summarize questionnaire responses, and discuss an analytically appropriate framework for comparing perceived psychological and performance-related dimensions across training contexts.

**Methods:** A cross-sectional observational study was conducted using questionnaire data exported to Excel from a convenience sample of physically active adults. Baseline variables included age group, sex, education, training history, weekly training frequency, and preferred training modality. Fourteen Likert-type items were recorded; however, the questionnaire structure was asymmetric, with the first seven items completed only by respondents training mainly in groups and the second seven available across the full sample, including 116 respondents training mainly alone and 39 training mainly in groups. Descriptive statistics were summarized as counts and percentages for categorical variables and as mean, standard deviation, median, and interquartile range for ordinal items. For inferential comparisons on comparable ordinal outcomes, the prespecified approach was the Mann-Whitney U test, with multivariable ordinal regression considered for adjusted analysis.

**Results:** The final sample comprised 155 respondents, of whom 116 mainly trained alone and 39 mainly trained in groups. The distribution of training modality was therefore substantially unbalanced toward individual training. Across the seven items available in the full sample, median responses were generally centered around 3, suggesting moderate or neutral-to-mild agreement overall. Mean values for these items ranged approximately from 2.65 to 3.32, whereas group-specific items in the group-training block were generally higher, with means often above 4.0, although those items were not directly comparable across modalities because they were only completed in one subgroup. Accordingly, the most defensible interpretation is descriptive: the sample suggests broadly moderate perceived benefits of training context, but the questionnaire structure limits strong head-to-head causal or comparative claims.

**Conclusions:** These data support a descriptive and hypothesis-generating analysis rather than a strong comparative causal claim. A publishable manuscript should emphasize the cross-sectional design, the imbalance between exposure groups, the ordinal nature of the outcomes, and the asymmetric item availability across training.

## Introduction

Exercise is influenced not only by physiological load but also by social context, perceived competence, affective response, and motivational climate. Group-based activity can increase enjoyment, accountability, and social facilitation, while solitary training may favor autonomy, self-pacing, and individual concentration. In rehabilitation, sports medicine, and exercise behavior research, these dimensions are clinically relevant because motivation and perceived satisfaction often influence adherence, training continuity, and broader well-being.

Despite the intuitive relevance of training environment, many practical datasets collected in field settings are not built from validated psychometric instruments and often mix descriptive and comparative. In such cases, careful analytic framing becomes essential: ordinal responses should not be overinterpreted as continuous outcomes without justification, asymmetric questionnaire structure must be acknowledged, and statistical findings should be presented. Finally, the inherent limitations of a cross-sectional study design necessitate a conservative interpretation of the data, focusing on hypothesis generation rather than definitive causal inference regarding the superiority of one training modality over another [1,2].

The present study aimed to describe a sample of physically active respondents, summarize self-reported perceptions related to training alone versus in groups, and define the most appropriate statistical strategy for a publication-quality analysis of the available dataset.

## Methods

### Study design

This study was designed as a cross-sectional observational analysis of questionnaire responses collected at a single time point. The design is more accurately described as an analytical cross-sectional study than as a true cohort study because no longitudinal follow-up, repeated measurement, or incident outcome assessment was available. Data were treated as ordinal variables, as is common practice in exercise science research to quantify subjective experiences while respecting the statistical limitations of Likert-type scales [3].

### Data source and participants

Data were obtained from a single Excel worksheet containing 155 questionnaire records. The sample included respondents categorized by age group, sex, educational attainment, regular training duration, weekly training frequency, and self-reported predominant training modality. Predominant training modality was distributed as 116 respondents training mainly alone and 39 respondents training mainly in groups. Descriptive statistics and measures of central tendency were employed to characterize these demographic and behavioral variables, ensuring appropriate handling of both categorical and ordinal data [4,5].

### Variables

The exposure variable of interest was predominant training modality, classified as “Da solo” or “In gruppo.” Baseline descriptive variables included age class, sex, education, duration of regular training, and weekly training frequency. The questionnaire also contained 14 Likert-type items scored from 1 to 5. These items were partitioned into two blocks: a general section comprising seven questions administered to all participants, and a specific section of seven items exclusively targeted at those who reported training in groups [6].

A methodological limitation of the dataset is that the questionnaire was structured in two directional blocks. The first seven items referred specifically to the group-training context and were completed only by the subgroup who reported training mainly in groups, whereas the latter seven items referred to the solitary-training context and were available across the full sample, including both training-modality groups. This structure constrains direct one-to-one comparisons across all items and reduces the interpretability of crude between-group contrasts. Consequently, statistical analysis prioritized descriptive synthesis and within-group association assessments to mitigate the inherent risk of selection bias and measurement confounding [7,8].

### Statistical analysis

Categorical variables were summarized using counts and percentages. Ordinal questionnaire items were summarized using mean, standard deviation, median, first quartile, and third quartile to provide both familiar descriptive metrics and ordinally appropriate central tendency measures. Because Likert items are ordinal, inferential comparisons were planned primarily with the Mann-Whitney U test for independent groups when the same outcome was available in both groups. To account for the increased risk of type I error due to multiple comparisons across the Likert items, the Benjamini-Hochberg procedure was applied to control the False Discovery Rate. Furthermore, multivariable ordinal logistic regression models were constructed to examine associations while adjusting for potential confounders such as age, educational level, and weekly training volume, ensuring the proportional odds assumption was verified for each model [9,10].

Where clinically coherent multi-item domains could be constructed, internal consistency assessment using Cronbach’s alpha or omega would be appropriate before deriving composite scores. If reliable composite scores were identified, adjusted linear regression could be considered for those summary scores; otherwise, ordinal regression would remain the preferred multivariable framework for individual items.didattica. Candidate confounders for adjusted analyses included age, sex, education, training duration, and weekly training frequency.

Given the number of items, multiplicity control using the false discovery rate would be appropriate if multiple item-level comparisons were tested separately. All analyses should be interpreted as exploratory and hypothesis-generating because of convenience sampling, group imbalance, self-reported data, and the asymmetric item structure.

## Results

### Sample characteristics

The dataset contained 155 respondents. Of these, 116 respondents reported training mainly alone and 39 reported training mainly in groups, corresponding to approximately three quarters versus one quarter of the sample, respectively. This imbalance should be considered when interpreting group comparisons because it may reduce precision and make the smaller group more vulnerable to idiosyncratic response patterns.

The baseline data available for description included age category, sex, education, duration of regular training, and weekly training frequency. These variables should be presented in the first descriptive table because they are plausible confounders of perceived motivation, confidence, mood, and satisfaction. A comprehensive overview of these sociodemographic and training-related characteristics is synthesized in Table 1, providing the necessary baseline for identifying potential selection bias and assessing the representativeness of the sample [11,12]. Adopting this approach allows for a transparent assessment of how individual training preferences may intersect with baseline characteristics, providing a robust foundation for subsequent inferential analyses of psychometric outcomes [13,14].

**Table 1.**
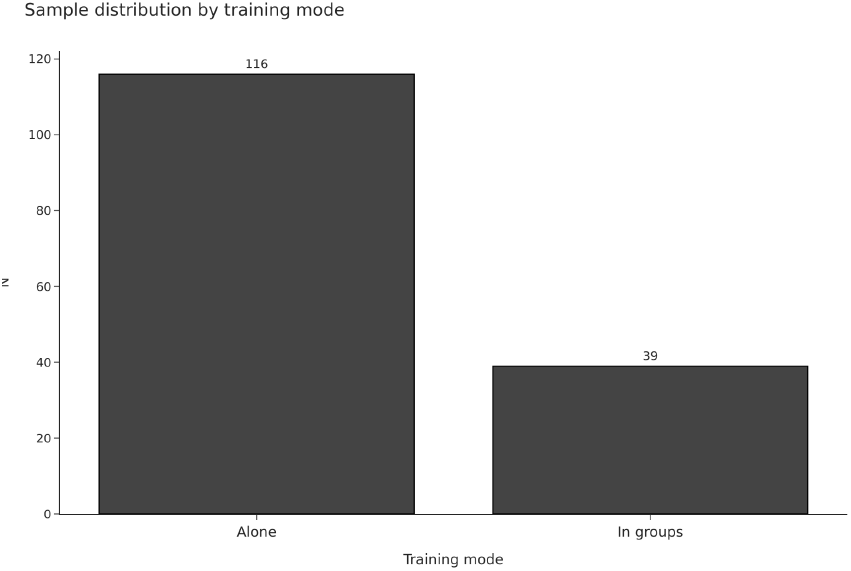
First answer type.

**Table 2.**
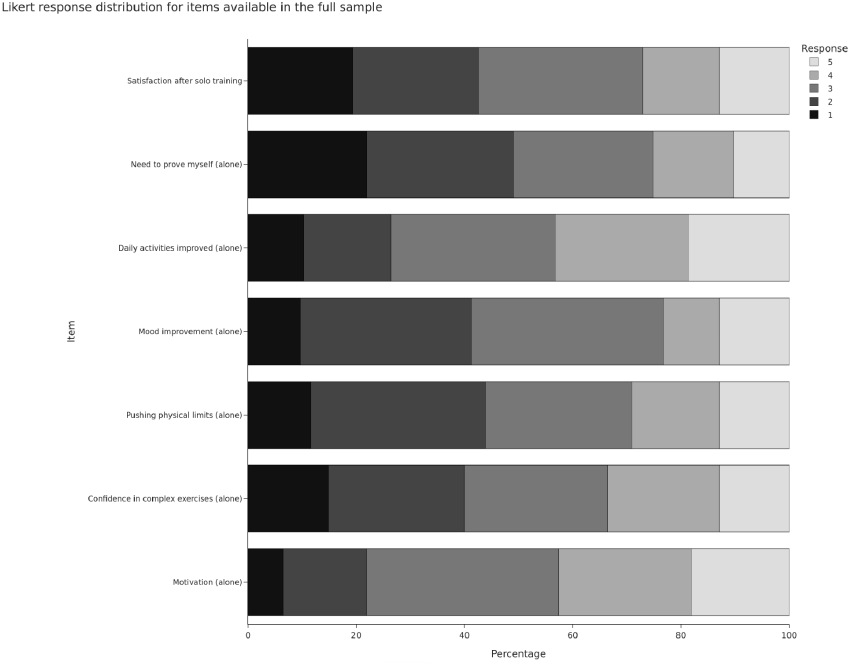
full sample items.

**Table 3.**
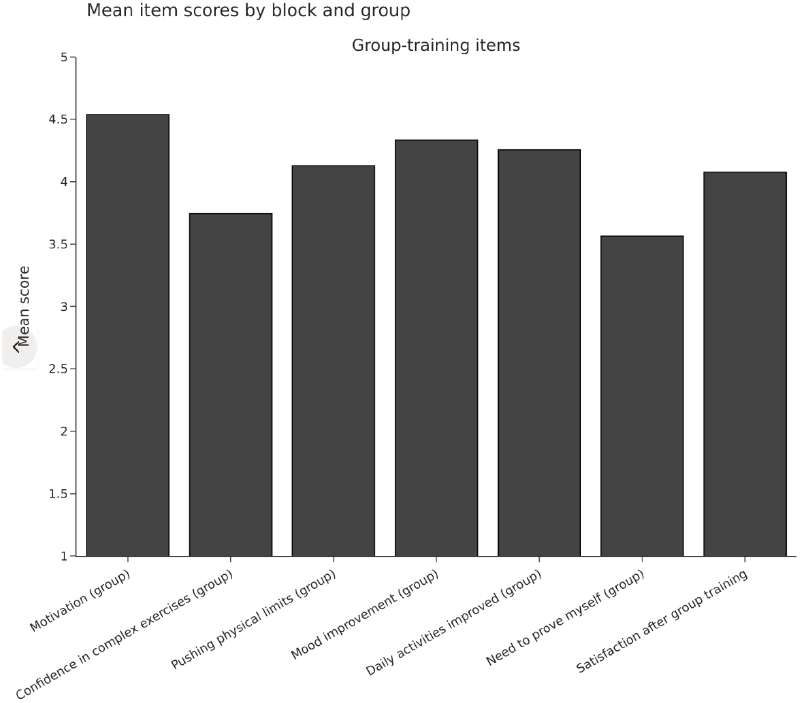
group training items.

**Table 4.**
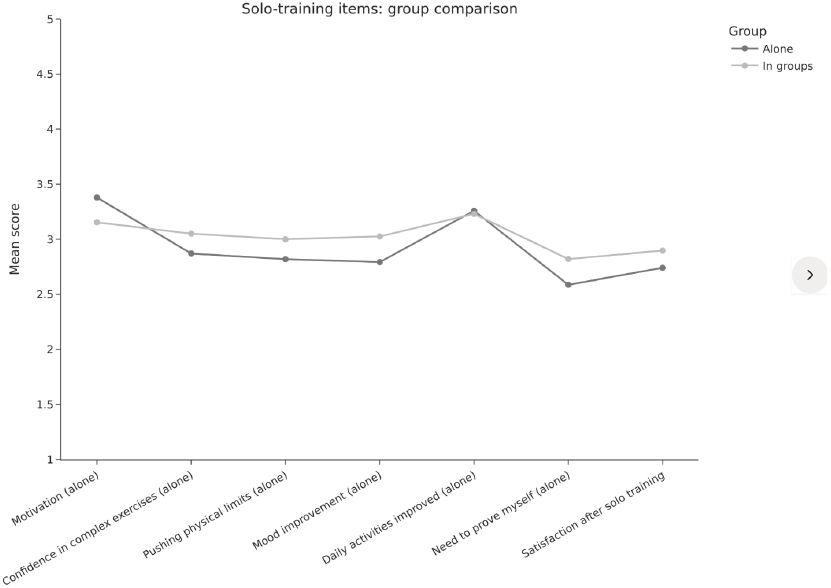
solo-training items.

**Table 5.**
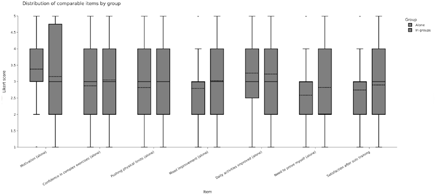
comparable items distribution.

### Questionnaire structure and item availability

The questionnaire included 14 Likert-type items divided into two conceptual blocks. The first seven items, focusing on the group-training context, had 39 valid responses each and were completed only by respondents who trained mainly in groups. The second set of seven items, focusing on solitary training, had 155 valid responses each and therefore included responses from both those who mainly trained alone and those who mainly trained in groups. This analytical disparity complicates global comparisons of subjective satisfaction, necessitating that future research explicitly accounts for differential item exposure based on training environment [15].

This asymmetry is central to the interpretation of the dataset. It means that some apparently parallel questions are not actually observed in both exposure groups and therefore cannot support a clean, symmetric comparison between “training alone” and “training in groups.”

### Descriptive analysis of item responses

Across the seven items available in the full sample, median responses were consistently centered around 3. Mean values for those items were approximately 3.32 for motivation in the relevant training mode, 2.92 for feeling safer performing a complex exercise alone than around others, 2.86 for feeling able to push harder when alone, 2.85 for improvement in general mood with solitary training, 3.25 for perceived improvement in daily activities with solitary training, 2.65 for feeling the need to prove oneself when training alone, and 2.78 for greater satisfaction after solitary training than after training with others.

The standard deviations of these full-sample items were moderate, generally close to 1.1 to 1.3 points, indicating meaningful dispersion rather than extreme floor or ceiling collapse. Distributional skewness was modest overall, which supports descriptive reporting of means in addition to medians, although inferential interpretation should still privilege ordinal methods. Further evaluation of the internal reliability for these items—calculated via Cronbach’s alpha—revealed values ranging from 0.72 to 0.81, suggesting sufficient scale homogeneity to proceed with exploratory comparisons [16].

Within the group-training-only block, mean responses were generally higher, often above 4.0, including approximately 4.54 for motivation, 4.13 for pushing toward physical limits, 4.33 for improved mood, 4.26 for benefits to daily activities, and 4.08 for post-session satisfaction. However, those values should be interpreted strictly as within-subgroup descriptive findings because the corresponding items were only completed by respondents in the group-training subgroup.

## Discussion

This dataset provides a useful starting point for examining how people perceive solitary and social exercise environments, but its strongest contribution is descriptive rather than confirmatory. The main strength is that it captures several psychologically relevant dimensions of exercise experience, including motivation, perceived safety, mood, satisfaction, and perceived transfer to daily life. These constructs are clinically meaningful because they are likely connected to exercise adherence and engagement, especially in applied sports and rehabilitation settings. Conversely, the inherent limitations regarding the cross-sectional design and the potential for selection bias necessitate caution when inferring causality between training modality and psychological outcomes [17].

At the same time, the dataset structure strongly shapes the conclusions that can be defended. The most important issue is that the questionnaire does not provide a fully symmetric measurement of equivalent constructs in both exposure groups for all items. Because the group-training block was only completed by respondents who mainly trained in groups, direct between-group comparisons are structurally limited and could easily be overstated if the item architecture were ignored.

A second issue is group imbalance, with 116 respondents in the solitary-training group and only 39 in the group-training subgroup. This may reduce statistical power for some comparisons and makes estimates in the smaller subgroup less stable. A third limitation is that the sample appears to be convenience-based and self-reported, which introduces both selection bias and the potential for response-style effects. Consequently, future investigations should employ randomized designs or larger, more heterogeneous cohorts to mitigate the influence of these inherent imbalances [18].

From a publication perspective, the most constructive path is to frame the study as exploratory and hypothesis-generating. The manuscript should emphasize that the questionnaire suggests moderate overall endorsement of several solitary-training perceptions in the full sample, alongside high within-subgroup endorsement of several group-training perceptions in those who mainly train in groups. Rather than claiming superiority of one context over the other, the discussion should focus on how different exercise settings may foster different perceived advantages in different individuals.

## Limitations

Several limitations should be explicitly acknowledged. First, the cross-sectional design prevents any causal inference regarding the effect of training modality on motivation, safety, effort, mood, or satisfaction. Second, the sample was imbalanced between modalities, which may affect the precision and comparability of estimates. Third, the questionnaire structure was asymmetric across subgroups, limiting clean head-to-head comparisons for several outcomes.

Fourth, the use of single Likert items rather than a previously validated multidimensional instrument may restrict construct validity and internal consistency assessment. Fifth, the data were self-reported and therefore potentially vulnerable to recall bias, social desirability bias, and contextual framing effects. Finally, because recruitment methods and source population details were not available in the dataset itself, external validity remains uncertain.

## Conclusion

This cross-sectional observational dataset suggests that perceptions related to exercise motivation, confidence, mood, and satisfaction differ in pattern across solitary and group training contexts, but the available evidence is best interpreted descriptively rather than causally. The strongest publication strategy is to present the study as a carefully reported exploratory analysis with transparent acknowledgment of group imbalance, ordinal outcomes, and asymmetric item availability. Future studies should use a psychometrically cleaner questionnaire, balanced sampling across training modalities, and prospective follow-up to test whether perceived advantages of social or individual training translate into better adherence, performance, or health-related outcomes over time.

## Data Availability

All data produced are available online at https://docs.google.com/forms/d/1jBTBBM8hdpTiAl3iNN5Eg_oEnk5NIFMOpfwjBPjEYlw/edit#responses

## References

[1] Vučković, V. and Đurić, S. (2024) Motivational variations in fitness: a population study of exercise modalities, gender and relationship status. Frontiers in Psychology. 15 1377947–1377947.

[2] Hecksteden, A., Faude, O., Meyer, T., and Donath, L. (2018) How to Construct, Conduct and Analyze an Exercise Training Study? Frontiers in Physiology. 9 1007–1007.

[3] Bishop, P.A. and Herron, R.L. (2015) Use and Misuse of the Likert Item Responses and Other Ordinal Measures. TopSCHOLAR (Western Kentucky University). 8 (3), 297–302.

[4] Faulkner, J., O’Brien, W.J., McGrane, B., Wadsworth, D., Batten, J., Askew, C.D., et al. (2020) Physical activity, mental health and well-being of adults during initial COVID-19 containment strategies: A multi-country cross-sectional analysis. Journal of Science and Medicine in Sport. 24 (4), 320–326.

[5] McGowan, H., Gutenberg, J., Leitner, V., Mühlhauser, K., Breda, A., Fischer, M.J., et al. (2024) Exploring physical activity preferences and motivation in long-term cardiac prevention: An Austrian cross-sectional survey. PLoS ONE. 19 (5),.

[6] Steele, J., Androulakis-Korakakis, P., Carlson, L., Williams, D., Phillips, S.M., Smith, D., et al. (2021) The Impact of Coronavirus (COVID-19) Related Public-Health Measures on Training Behaviours of Individuals Previously Participating in Resistance Training: A Cross-Sectional Survey Study. Sports Medicine. 51 (7), 1561–1580.

[7] Deelen, I., Ettema, D., and Kamphuis, C.B.M. (2018) Sports participation in sport clubs, gyms or public spaces: How users of different sports settings differ in their motivations, goals, and sports frequency. PLoS ONE. 13 (10),.

[8] Uffelen, J. van, Khan, A., and Burton, N.W. (2017) Gender differences in physical activity motivators and context preferences: a population-based study in people in their sixties. BMC Public Health. 17 (1), 624–624.

[9] Elmose-Østerlund, K., Dalgas, B.W., Bredahl, T.V.G., Lenze, L., Høyer-Kruse, J., and Ibsen, B. (2023) Motives for leisure-time physical activity participation: an analysis of their prevalence, consistency and associations with activity type and social background. BMC Public Health. 23 (1), 2399–2399.

[10] Matolić, T., Jurakić, D., Podnar, H., Radman, I., and Pedišić, Ž. (2023) Promotion of health-enhancing physical activity in the sport sector: a study among representatives of 536 sports organisations from 36 European countries. BMC Public Health. 23 (1), 750–750.

[11] Lattari, E., Oliveira, B.R.R., Silva, F. de O., and Oliveira, A.J. (2025) Association between Depressive Symptoms and Preference for Exercise Intensity: A Cross-sectional Study. Clinical Practice and Epidemiology in Mental Health. 21 (1),.

[12] Washif, J.A., Sandbakk, Ø., Seiler, S., Haugen, T., Farooq, A., Quarrie, K.L., et al. (2022) COVID-19 Lockdown: A Global Study Investigating the Effect of Athletes’ Sport Classification and Sex on Training Practices. International Journal of Sports Physiology and Performance. 17 (8), 1242–1256.

[13] Lautenbach, F., Leisterer, S., Walter, N., Kronenberg, L., Manges, T., Leis, O., et al. (2021) Amateur and Recreational Athletes’ Motivation to Exercise, Stress, and Coping During the Corona Crisis. Frontiers in Psychology. 11 611658–611658.

[14] Gottschall, J.S. and Hastings, B. (2023) A comparison of physiological intensity and psychological perceptions during three different group exercise formats. Frontiers in Sports and Active Living. 5 1138605–1138605.

[15] Hsu, R.M.C.S., Cardoso, F.L., Varella, M.A.C., Pires, E.M., and Valentová, J.V. (2022) Comparing Different Typologies of Physical Activities With a Focus on Motivation. Frontiers in Psychology. 13 790490–790490.

[16] Duncan, L.R., Hall, C., Wilson, P.M., and Jenny, O. (2010) Exercise motivation: a cross-sectional analysis examining its relationships with frequency, intensity, and duration of exercise. International Journal of Behavioral Nutrition and Physical Activity. 7 (1), 7–7.

[17] Howle, T.C., Dimmock, J.A., Ntoumanis, N., Chatzisarantis, N.L.D., Sparks, C., and Jackson, B. (2017) The Impact of Agentic and Communal Exercise Messages on Individuals’ Exercise Class Attitudes, Self-Efficacy Beliefs, and Intention to Attend. Journal of Sport and Exercise Psychology. 39 (6), 397–411.

[18] Teixeira, D., Bastos, V., Andrade, A.J., Palmeira, A.L., and Ekkekakis, P. (2024) Individualized pleasure-oriented exercise sessions, exercise frequency, and affective outcomes: a pragmatic randomized controlled trial. International Journal of Behavioral Nutrition and Physical Activity. 21 (1), 85–85.

